# eoPred: Predicting the placental phenotype of early-onset preeclampsia using DNA methylation

**DOI:** 10.1101/2023.05.17.23290125

**Authors:** I. Fernández-Boyano, A.M. Inkster, V. Yuan, W.P. Robinson

**Affiliations:** University of British Columbia

## Abstract

**Background:** A growing body of literature has reported molecular and histological changes in the human placenta in association with preeclampsia (PE). Placental DNA methylation (DNAme) and transcriptomic patterns have revealed molecular subgroups of PE that are associated with placental histopathology and clinical phenotypes of the disease. However, the heterogeneity of PE both across and within subtypes, whether defined clinically or molecularly, complicates the study of this disease. PE is most strongly associated with placental pathology and adverse fetal and maternal outcomes when it develops early in pregnancy. We focused on placentae from pregnancies affected by preeclampsia that were delivered before 34 weeks of gestation to develop eoPred, a predictor of the DNAme signature associated with the placental phenotype of early-onset preeclampsia (EOPE).

**Results:** Public data from 83 placental samples (HM450K), consisting of 42 EOPE and 41 normotensive preterm birth (nPTB) cases, was used to develop eoPred - a supervised model that relies on a highly discriminative 45 CpG DNAme signature of EOPE in the placenta. The performance of eoPred was assessed using cross-validation (AUC=0.95) and tested in an independent validation cohort (n=49, AUC=0.725). A subset of fetal growth restriction (FGR) and late-PE cases showed a similar DNAme profile at the 45 predictive CpGs, consistent with the overlap in placental pathology between these conditions. The relationship between the EOPE probability generated by eoPred and various phenotypic variables was also assessed, revealing that it is associated with gestational age, and it is not driven by cell composition differences.

**Conclusions:** eoPred relies on a 45 CpG DNAme signature to predict EOPE, and it can be used in a discrete or continuous manner. Using this classifier should 1) improve the consistency of future placental DNAme studies of PE and placental insufficiency, 2) facilitate identifying cases of EOPE in public data sets and 3) importantly, standardize the placental diagnosis to allow better cross-cohort comparisons. Lastly, classification of cases with eoPred should be useful for testing associations between placental pathology and genetic or environmental variables.

## Background

Preeclampsia (PE) is a hypertensive disorder of pregnancy, and is one of the leading causes of maternal and perinatal morbidity and mortality worldwide. The Society of Obstetricians and Gynecologists of Canada defines PE as gestational hypertension with new-onset proteinuria or one or more adverse conditions, which include maternal end-organ complications and evidence of uteroplacental dysfunction (Magee et al., 2014). While the global incidence of PE is estimated as 4.6% (Abalos et al., 2013), this is an approximation limited by the lack of data from many regions, and varies by population, race, and socioeconomic status (Silva et al., 2008; Lisonkova and Joseph, 2013; Wang et al., 2021). Encompassing a diverse range of clinical presentations and outcomes, PE is a complex and multifactorial disease, which exists on a spectrum of severity. Among others, chronic hypertension and prior PE are well-known risk factors for PE (Bartsch et al., 2016). Placental, maternal, and paternal genetics also significantly contribute to the pathogenesis of PE (Williams and Broughton Pipkin, 2011; Wang et al., 2022). The combination of such genetic and environmental factors plays an important role in the development of PE, which is a two-stage process whereby placental dysfunction leads to maternal onset of disease (Redman et al., 2014); several studies have observed PE-associated molecular and histopathological changes in the human placenta, likely reflecting the observed placental dysfunction (Leavey et al., 2016; Benton et al., 2018; Wilson et al., 2018). Characterization of placental PE-associated changes can complement clinical findings to gain a better understanding of the disease and assist with its classification into more homogeneous subtypes.

The umbrella syndrome of PE is likely the junction where at least two distinct, and likely interacting, pathways to disease converge, initially coined as “placental” and “maternal” (Roberts and Escudero, 2012; Redman et al., 2014; Redman, 2017; Staff and Redman, 2018; Staff, 2019, 201). Clinically, diagnosis before or at/after 34 weeks gestational age (GA) may be used to define two subtypes of PE: early-onset PE (EOPE) and late-onset PE (LOPE) (Brown et al., 2018). EOPE, which often has a greater placental involvement (Redman, 2017), is more severe, frequently overlaps with other pathologies such as fetal growth restriction (FGR), and presents with placental pathology such as villous infarctions and hypermaturation. In contrast, LOPE may be more heavily influenced by predisposing maternal factors despite normal placentation, and is often milder (Staff, 2019). EOPE is thought to originate with reduced placental perfusion because of incomplete vascular remodelling during placentation (Staff, 2019). Oxidative stress secondary to malperfusion then leads to trophoblast damage, which eventually results in the systemic inflammatory response and ensuing hypertensive syndrome (Roberts and Hubel, 2009; Staff, 2019), although the mechanisms linking the two stages are still being elucidated (Roberts and Hubel, 2009). In contrast, the development of LOPE does not necessitate inadequate placentation but instead may result from compression of the terminal villi at term, which impedes appropriate perfusion, leading to syncytiotrophoblast hypoxia as in EOPE (Staff, 2019). All PE is thus associated with placental syncytiotrophoblast stress (Staff, 2019), albeit with distinct underlying causes and timing. Maternal factors such as obesity also contribute to all stages of the disease and play an important role in disease severity.

In the human placenta, DNA methylation (DNAme) has a unique profile that shifts throughout gestation in association with changes in cell composition, gene expression, and in response to pregnancy complications, among other factors (Robinson and Price, 2015; Yuan et al., 2021). Many studies have reported PE-associated alterations in placental DNAme (Blair et al., 2013; Chu et al., 2014; Martin et al., 2015; Yeung et al., 2016; Kim, 2017; Wilson et al., 2018; Wang et al., 2019a; Lim et al., 2020; Workalemahu Tsegaselassie et al., 2020), but findings are not consistently reproduced across studies (Cirkovic et al., 2020). Discrepancies between studies in definitions of PE used, study design (e.g., analysing all PE compared to analysing specific subtypes such as EOPE, LOPE), the use of different metrics to assess reproducibility (e.g., a CpG found to be significantly differentially methylated in one cohort may not replicate in an independent cohort, but a proximal and correlated CpG might), and platforms used to measure DNAme, among others, can contribute to poor reproducibility across studies. Identification of homogeneous subtypes of PE is essential to successfully identify and manage each subtype (Myatt et al., 2014).

PE-associated molecular variation in the placenta may be used to refine current classification of PE. Wilson et al. found that the placental DNAme profile of intermediate onset PE (<36 weeks) co-occurring with FGR is similar to that of EOPE, suggesting that the clinical threshold of 34 weeks is imperfect and will misclassify cases (Wilson et al., 2018). Further, integration of DNAme with transcriptional information revealed up to four molecular PE subtypes (Leavey et al., 2016). Additionally, several histopathological findings including maternal vascular malperfusion lesions were found to associate with each molecular PE subcluster (Leavey et al., 2016). Severity also existed on a gradient within each subcluster, supporting the hypothesized connection between molecular changes and pathology in the placenta, as well as illustrating the value of studying DNAme and transcriptomic profiles to characterize PE.

To address these challenges, we developed eoPred, a DNAme-based model that uses 450K Illumina DNAme microarray data from placental chorionic villus samples taken at delivery to find a placental signature of EOPE. This model was validated by predicting the disease status of samples in an independent cohort. Furthermore, the model outputs a continuous probability that can be valuable in correlating placental changes with environmental and genetic variables, as well as with postnatal outcomes.

## Methods

### Study data

Placental DNAme data (n=401) were collected from eight Illumina Infinium HumanMethylation450 Beadchip array (HM450K) datasets (GSE100197 (Leavey et al., 2018; Wilson et al., 2018), GSE103253 (Herzog et al., 2017), GSE120981 (Monteagudo-Sánchez et al., 2019), GSE73375 (Martin et al., 2015), GSE75196 (Yeung et al., 2016), GSE98224 (Leavey et al., 2018; Wilson et al., 2018), GSE49343 (Blair et al., 2014)) and one Illumina Infinium HumanMethylation850 (EPIC) dataset (GSE203396 (Inkster et al., 2022) available on Gene Expression Omnibus (GEO). Using samples from these datasets, we created three separate groups for i) model training, ii) model validation and iii) model exploration (Table 1). To develop a predictive model for canonical EOPE, the training group was restricted to EOPE and normotensive preterm birth (nPTB) samples to avoid confounding by inclusion of LOPE or normotensive fetal growth restriction (FGR), which are heterogeneous groups that may partly overlap with the placental phenotype of EOPE, and normotensive term samples, which may differ from EOPE due to GA-associated changes rather than due to pathology. We chose to generate a binary classifier that differentiated between EOPE and nPTB rather than a multi-class predictor (i.e., also including FGR and LOPE) because of the clear placental dysfunction observed in EOPE compared to other pathologies. The validation group was also comprised of EOPE and nPTB samples. The exploratory group consisted of a broader mix of samples with different pathologies, exclusive of those used in the training and validation groups, to better understand the application of our model to a variety of placental phenotypes. Cord blood DNAme data was downloaded from GSE110829 to test the tissue-specificity of eoPred.

**Table 1.**
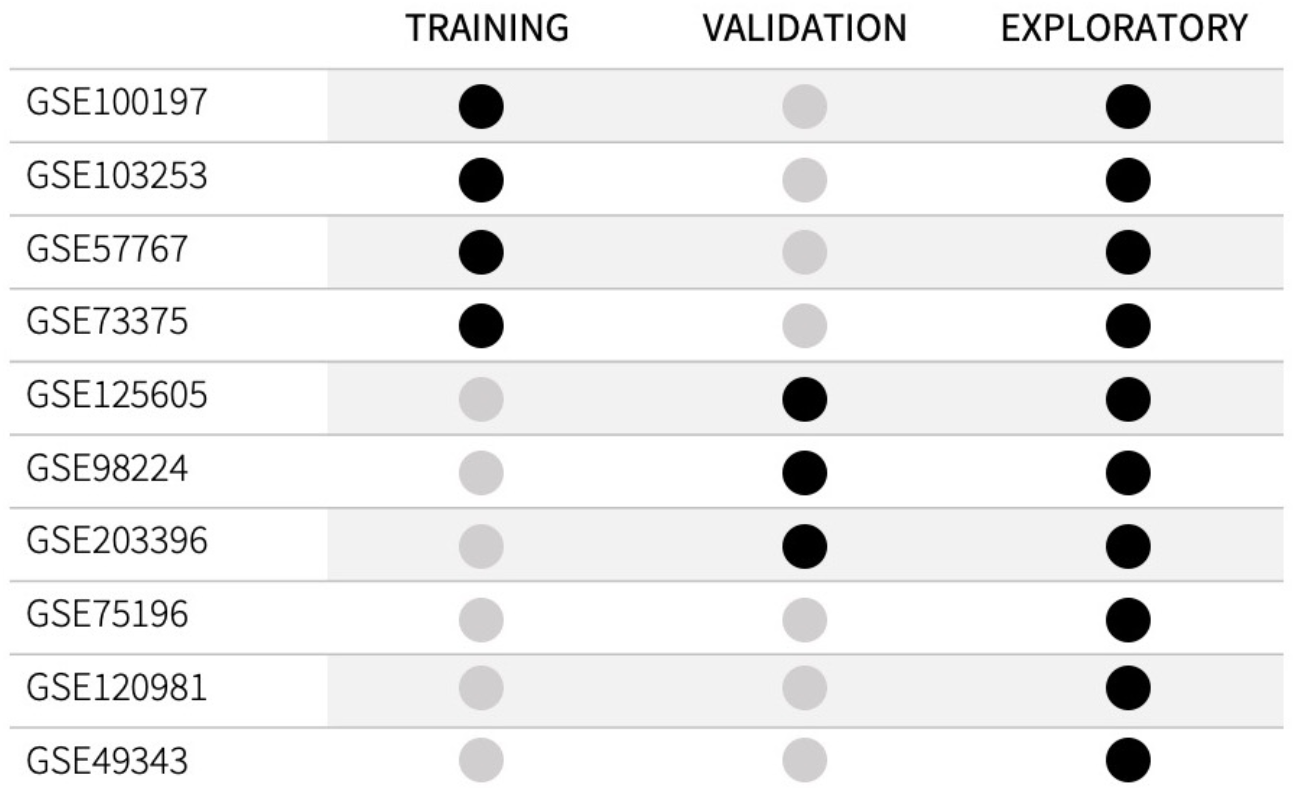
Arrangement of datasets from the Gene Expression Omnibus into the three study groups. Samples in the training and validation groups only included those samples labelled as EOPE and nPTB by dataset authors. Samples from GSE100197, GSE103253, GSE57767, GSE73375, GSE125605, GSE98224, and GSE203396 that were not included in the training and validation groups (i.e., from etiologies other than EOPE or nPTB) are in the exploratory group.

The training group (n=89) was assembled with the criteria that each of the datasets included (GSE100197, GSE103253, GSE57767, GSE73375) must contain a mixture of EOPE (n=47) and nPTB (n=42) cases, balanced by dataset (i.e., if a dataset included only 1 EOPE sample and several nPTB samples, it was excluded). The validation group (n=49) was selected from three datasets (GSE125605, GSE98224, GSE203396) with a total of 38 EOPE and 11 nPTB samples. An exploratory group (n=329) was then constructed from all remaining samples (GSE100197, GSE103253, GSE57767, GSE73375, GSE125605, GSE98224, GSE120981, GSE75196, GSE49343, GSE203396) that had a diagnosis other than EOPE or nPTB (i.e., late-PE, nFGR, CPM16 and nTB). This third group was assembled separately from the validation group to avoid any bias that might result from processing the data in the validation group with a dataset that was also included in the training cohort, even if the specific samples were not actually used for training the model. Demographic characteristics of each group are summarized in Table 2.

**Table 2.** Demographic characteristics of each constructed group included in this study. The data reflects the number of samples after processing.

|  | Training Cohort N (%) | Validation Cohort N (%) | Exploratory Cohort N (%) | GSE110829 N (%) |
| --- | --- | --- | --- | --- |
| Early-onset preeclampsia (EOPE) | 42 (51%) | 38 (78%) | 2 (1%) | 10 (9%) |
| Gestational age range (weeks) | 31.6 (25-37) | 33.25 (26-39) | 32.78 (32-33) | 29.91 (26-33) |
| Placental sex (XY/XX) | 23/19 | 16/22 | 1/1 | 2/8 |
| Late preeclampsia (Late-PE) | 0 | 0 | 86 (26%) | 10 (9%) |
| Gestational age range (weeks) | NA | NA | 36.97 (34-41) | 35.57 (34-38) |
| Placental sex (XY/XX) | NA | NA | 45/41 | 3/7 |
| Fetal growth restriction (FGR) | 0 | 0 | 42 (14%) | 0 |
| Gestational age range (weeks) | NA | NA | 36.43 (28-40) | NA |
| Placental sex (XY/XX) | NA | NA | 24/22 | NA |
| Normotensive preterm birth (nPTB) | 41 (49%) | 11 (22%) | 3 (1%) | 47 (43%) |
| Gestational age range (weeks) | 33.76 (25-36) | 34.42 (31-36) | 33.11 (28-37) | 30.44 (23-37) |
| Placental sex (XY/XX) | 23/18 | 5/6 | 2/1 | 29/18 |
| Normotensive term birth (nTB) | 0 | 0 | 181 (55%) | 43 (39%) |
| Gestational age range (weeks) | NA | NA | 38.62 (37-42) | 38.58 (37-41) |
| Placental sex (XY/XX) | NA | NA | 92/89 | 19/24 |
| Trisomy 16 confined to the placenta (CPM16) | 0 | 0 | 10 (3%) | 0 |
| Gestational age range (weeks) | NA | NA | 34.10 (25-39) | NA |
| Placental sex (XY/XX) | NA | NA | 5/5 | NA |

As sub-classification of PE into EOPE and LOPE was not always reported by dataset authors for all samples, PE was classified as follows: samples reported as “PE”, were labelled as EOPE if delivered prior to 34 weeks and were otherwise classified as “late-PE”. No EOPE-reported cases were labelled as late-PE using this approach, since cases delivered prior to 34 weeks would by necessity have been diagnosed before 34 weeks. While PE cases with clinical onset <34 weeks (EOPE) but delivery after 34 weeks would have been labelled as late-PE using this approach, none of these were included in model development and testing but were reserved only for exploratory analysis.

There were also inconsistencies in the reporting of GA across datasets. GA was provided in weeks and days by five datasets (GSE100197, GSE98224, GSE125605, GSE110829, GSE203396), two datasets reported weeks without days (GSE73375, GSE75196), one dataset (GSE57767) only distinguished between term and preterm samples, and two datasets (GSE103253, GSE120981) did not provide any metric of GA. To harmonize the GA metric across studies, placental epigenetic GA was calculated using the robust placental clock (RPC) (Lee et al., 2019) for all samples, using the R package planet (Lee et al., 2019; Yuan, 2023).The difference between reported and predicted GA in the datasets in which reported GA was available is summarized in Figure S1 and Table S1 (Supplementary Materials); reported and predicted GA were highly correlated in the training group (R=0.94) and there was a median absolute error of 4 days across all samples in the three groups. We note that the robust placental clock was trained on GSE100197, amongst other datasets. Using RPC-predicted rather than reported GA did not change the classification of samples into EOPE and late-PE; importantly, no cases clinically classified as LOPE by the authors were re-labelled as EOPE using predicted GA (Table S1, Supplementary Materials). Genetic ancestry and cell composition were also calculated for all samples using planet (Yuan et al., 2019, 2021; Yuan, 2023).

### Data processing

Data were downloaded as IDAT files wherever possible, otherwise methylated and unmethylated intensities were used. The training, validation, and exploratory group, and the cord blood dataset, were each processed independently using the same sample exclusion criteria, and probe filtering criteria, and the same normalization method, beta-mixture quantile (BMIQ) from the R package wateRmelon (Pidsley et al., 2013).

After normalization, samples were removed from downstream analyses if they failed any of the following checks: i) mean inter-array correlation <95% (Edgar et al., 2017) ii) discordance between reported sex and chromosomal sex as inferred by X and Y chromosome fluorescence intensities (Heiss and Just, 2018), or iii) a measure of sample contamination based on allelic ratios of single-nucleotide polymorphism (SNP) probes implemented in the ewastools package (Heiss and Just, 2018). CpG probes were removed if they had poor quality signal in more than 5% of samples (detection P value > 0.01 or bead count < 3) (Aryee et al., 2014) or targeted loci cross-hybridizing to multiple sites (Price et al., 2013; Zhou et al., 2017), overlapped polymorphisms in the genome (Zhou et al., 2017), mapped to the X and Y chromosomes (Bibikova et al., 2011), or were non-variable both in this dataset and were reported as non-variable probes in the placenta (Edgar et al., 2017). To allow future application of eoPred on data collected with the Infinium MethylationEPIC v1.0 Beadchip array (EPIC), probes were filtered to those in common between the HM450K and EPIC platforms prior to developing the model. Probe filtering was only applied to the training group; to ensure that all 45 CpGs were present when applying eoPred, the other groups were normalized and checked for sample quality, but not filtered for poor-quality probes.

Lastly, batch correction was applied to the training group only using ComBat with the R package sva (Edgar et al., 2017) to correct for dataset differences. A model matrix was created with variables of interest (condition, sex, GA, and European and Asian ancestry coordinates assigned to each sample by planet (Yuan, 2023)) to preserve their variation, as recommended in (Leek et al., 2012). No batch correction was applied to the validation, exploratory, or cord blood groups.

After processing, 341,281 CpGs and 83 samples remained in the training group. The validation group consisted of 49 samples, the exploratory group was composed of 329 samples, and the cord blood data was comprised of 110 samples. Demographic data of the three constructed groups is shown in Table 1.

### Model development

R Markdown source files and knitted reports for the analysis can be found on GitHub at https://github.com/iciarfernandez/eoPred/. The R package mixOmics was used for all steps in model development and assessment (Rohart et al., 2017). Other R packages used to calculate model metrics were cvms (Renbo, 2016), MLmetrics (Yan, 2017), and caret (Kuhn, 2008). Repeated (n=50) M-fold (n=3) cross-validation (CV) was used to develop and assess the performance of eoPred. Three folds were chosen to ensure that enough samples were present in each fold, given the relatively small sample size of the cohort. One fold was reserved to assess model performance, and the remaining two folds were used to train the model. This process was repeated 50 times, and each of the 50 performance estimations were averaged to produce a single estimation.

The model was trained on mean DNAme beta values, using sparse partial least squares discriminant analysis (sPLS-DA), which performs simultaneous dimensionality reduction and feature selection to find a molecular signature that can discriminate samples based on an outcome category (in this case, EOPE or nPTB). sPLS-DA has been previously shown to perform well on high-dimensional genomics data (Lê Cao et al., 2011). After the optimal parameters (number of components, and number of features per component) were selected during CV, a final model was fit to the entire training data.

Overall misclassification error rate, receiver operating characteristic and area under the curve (ROC-AUC), sensitivity, and Brier score were the metrics used to assess the performance of the final model. A permutation test was run where a model with randomly permuted labels was trained and compared to the final model. The stability of the predictive DNAme-signature was also assessed using the *perf* function of the mixOmics package, which computes the frequency at which each CpG is selected in each CV run, and uses this to evaluate each CpG’s stability (i.e., a CpG selected with low frequency across all CV runs may suggest that it is relevant to a certain split of the training data, but not to others) (Rohart et al., 2017).

### Model validation

To test the performance of eoPred, the predict function in mixOmics (Rohart et al., 2017) was used to assign a predicted class to each of the 48 samples in the validation group. The maximum prediction distance was chosen as it resulted in the lowest error rate during the CV process. Each sample is thus assigned two prediction distances (one per outcome category) and the predicted class is the outcome category with the largest predicted dummy value. Class probabilities were calculated from transformed prediction distances using the softmax function, such that each sample was assigned two estimated probabilities of being classified as either EOPE or nPTB, which sum to 1. Each sample was then predicted to be the class with the largest class probability.

### Relationship between DNAme at predictive CpG probes and gene expression

Placental gene expression data was downloaded for GSE75010 (Affymetrix Human Gene 1.0 ST Array) and GSE44711 (Illumina HumanHT-12 V4.0 Expression Beadchip). Gene expression data from GSE44711 (n=16) matching DNAme samples from GSE100197 (dataset in the training group) was used as our discovery group while samples from GSE75010 (n=92) were used for validation as they matched data from GSE98224 (dataset in the validation group). Co-methylated regions (CMRs) were constructed from the 341,281 CpGs in the training group using the CoMeBack package (Gatev et al., 2020) and a Spearman correlation cutoff of 0.3. CMRs containing the 45 predictive CpGs in eoPred were selected, and genes in those CMRs were tested for differential expression by disease status (EOPE/nPTB), adjusting for GA and sex. Genes were considered to replicate differential expression by disease status at nominal significance (*p* < 0.05) with a change in expression in the same direction as the discovery cohort.

### Biological characterization of predictive CpG probes

To understand whether the CpGs selected by the model as predictive were biologically relevant to the pathophysiology of preeclampsia, two approaches were taken. First, the R package missMethyl (Phipson et al., 2016) was used to conduct gene ontology (GO) enrichment analysis. Genes associated with the 341,281 CpGs that the model was trained on were used as the background set for enrichment. Biological process GO terms were considered significantly enriched at FDR < 0.05.

## Results

### Development of a supervised model using placental DNAme that predicts EOPE

Using sparse partial least discriminant analysis (sPLS-DA), we developed a placental DNAme-based classifier that selected EOPE-associated CpG sites in a training group and were able to use this signature to accurately predict the disease status of samples in an independent validation group.

Parameter tuning during CV selected the optimal number of components and features (CpGs) per component, with a component being constructed from a linear combination of the features. One component constructed from 90 CpGs resulted in the lowest average overall misclassification error rate (OER) across all CV folds. The optimal number of components is typically K-1, with K being the number of classes to be predicted by the model. Thus, as expected, introducing more components in the model did not significantly improve CV performance. While this model performed well on CV (OER=0.10, AUC=0.97), 35% of the 90 CpGs selected had low stability (frequency of selection across CV folds < 0.5). These CpGs also contributed less than those with higher stability to the model’s discriminative ability, suggesting that a simpler model with less CpGs may be equally successful in predicting the outcome of new samples. To test this, we developed six additional models by selecting the number of CpGs *a priori* (75, 60, 45, 30, 15, 5) and assessed their performance on CV.

Decreasing the number of CpGs did not significantly affect model performance as evaluated using two discrete classification metrics (OER, ROC-AUC) and one probability-based metric (Brier score) (Table 3). The mean OER value of the seven models with different numbers of CpGs was 0.11 (±0.01 standard deviation); we saw a slight but non-significant increase in OER as the number of CpGs decreased (Figure 1a, Table 3). For all seven models, we also assessed the area under the curve (AUC, Table 3) of the receiver operating characteristic (ROC, Figure 1b) curve, which informs about the trade-off between the true positive rate and the false positive rate. Once again, the difference between the seven models on this metric was not significant, and all seven showed outstanding discrimination with an AUC of 0.95 (Applied Logistic Regression, 3rd Edition, n.d.). Lastly, we computed the average Brier score across CV folds for each model, which measures the mean squared difference between the predicted probability assigned to the possible outcomes for a given sample, and its actual outcome. All seven models had an average Brier Score of 0.14 (Table 3), which confirmed that the predicted probability of most samples matched the true likelihood of the outcome of interest.

**Figure 1.**
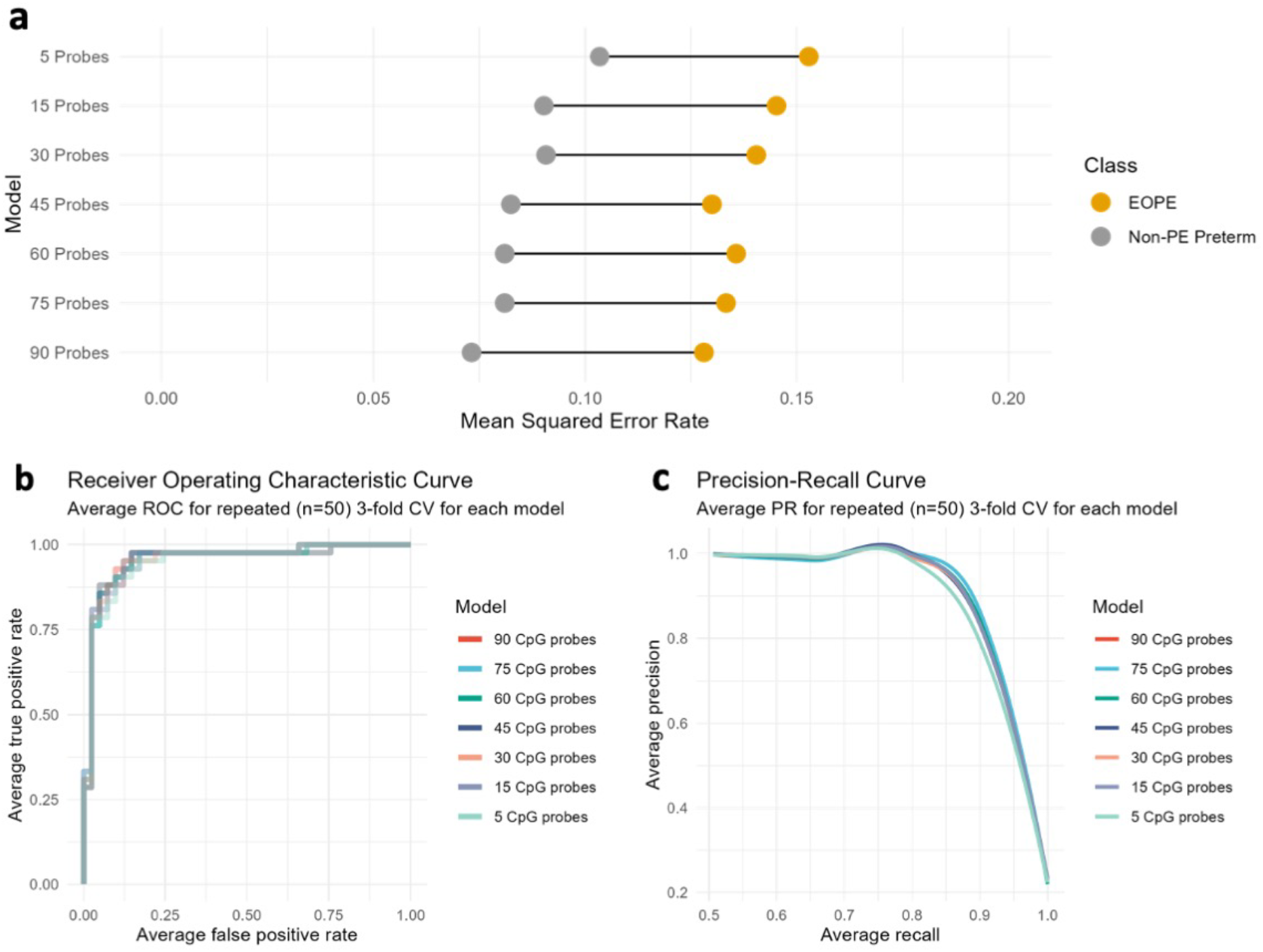
Cross-validation performance of the seven models based on varying number of CpGs. a) The performance of each model computed across CV folds according to the overall classification error rate (OER) of EOPE and nPTB samples. b) The receiver operating characteristic (ROC) curve of each model calculated across CV folds demonstrates a similar performance of the seven models. C) The precision-recall curve of each model calculated across CV folds, which also demonstrates a similar performance across all models.

**Table 3.** Average performance metrics from cross-validation folds in the seven models tested during development, and in the permuted model. OER is the overall error rate, defined as the number of all incorrect predictions divided by the total number of samples in the data. OER (EOPE) is calculated exclusively over the EOPE samples (n=38), and OER (nPTB) is calculated over the nPTB samples (n=11). AUC is the area under the receiver operating characteristic curve, it is a measure of separability. The Brier score measures the accuracy of probabilistic predictions.

| Model | OER | OER SD | OER (EOPE) | OER (nPTB) | AUC | Brier score |
| --- | --- | --- | --- | --- | --- | --- |
| 90 CpGs | 0.10 | 0.01 | 0.13 | 0.07 | 0.95 | 0.14 |
| 75 CpGs | 0.11 | 0.02 | 0.13 | 0.08 | 0.95 | 0.14 |
| 60 CpGs | 0.11 | 0.02 | 0.14 | 0.08 | 0.95 | 0.14 |
| 45 CpGs (Final Model) | 0.11 | 0.01 | 0.13 | 0.08 | 0.95 | 0.14 |
| 45 CpGs (Permuted Model) | 0.53 | 0.05 | 0.52 | 0.55 | 0.45 | 0.26 |
| 30 CpGs | 0.12 | 0.02 | 0.14 | 0.09 | 0.95 | 0.14 |
| 15 CpGs | 0.12 | 0.02 | 0.15 | 0.09 | 0.95 | 0.14 |
| 5 CpGs | 0.13 | 0.13 | 0.15 | 0.10 | 0.93 | 0.14 |

We selected the 45 CpG classifier as our final model for outcome prediction on independent data (Table S4, Supplementary Materials). While a model relying on a very small number of CpGs (e.g., 5) may demonstrate a good average performance on CV, it might not be as generalizable to new data. At the same time, the larger model (90 CpGs) initially selected by CV does not perform significantly better than any of the smaller models. Therefore, we chose a model with an intermediate number of features (45 CpGs) to evaluate on new data.

Lastly, we compared the average performance across CV folds of the final 45 CpG model to a 45 CpG model that was trained on randomly permuted labels. The goal of this permutation test was to assess whether our final model performs better than a model would be expected to by chance. Across all metrics, it was clear that the performance of the model with permuted labels was significantly worse (0.52 OER, AUC=0.45) as compared to our final model (Table 3).

### eoPred demonstrated good discriminatory ability (AUC=0.73) in an independent validation group

To test the performance of eoPred, we applied it to predict the disease status of 49 samples (38 EOPE, 11 nPTB) in the validation group. Each sample was assigned a probability of being classified as either EOPE or nPTB; while some were labelled with a high confidence, others were intermediate (Figure 2b). Youden’s J statistic, defined as the sum of sensitivity and specificity minus one, was used to choose an optimal threshold of 55% to classify samples (Figure 2a). However, this threshold can be modified by the user in the eoPred function, and it is encouraged that users explore the spread of probabilities in their data and determine a threshold that fits their question best. For example, higher probabilities of EOPE may be used to select samples with a more homogeneous placental phenotype.

**Figure 2.**
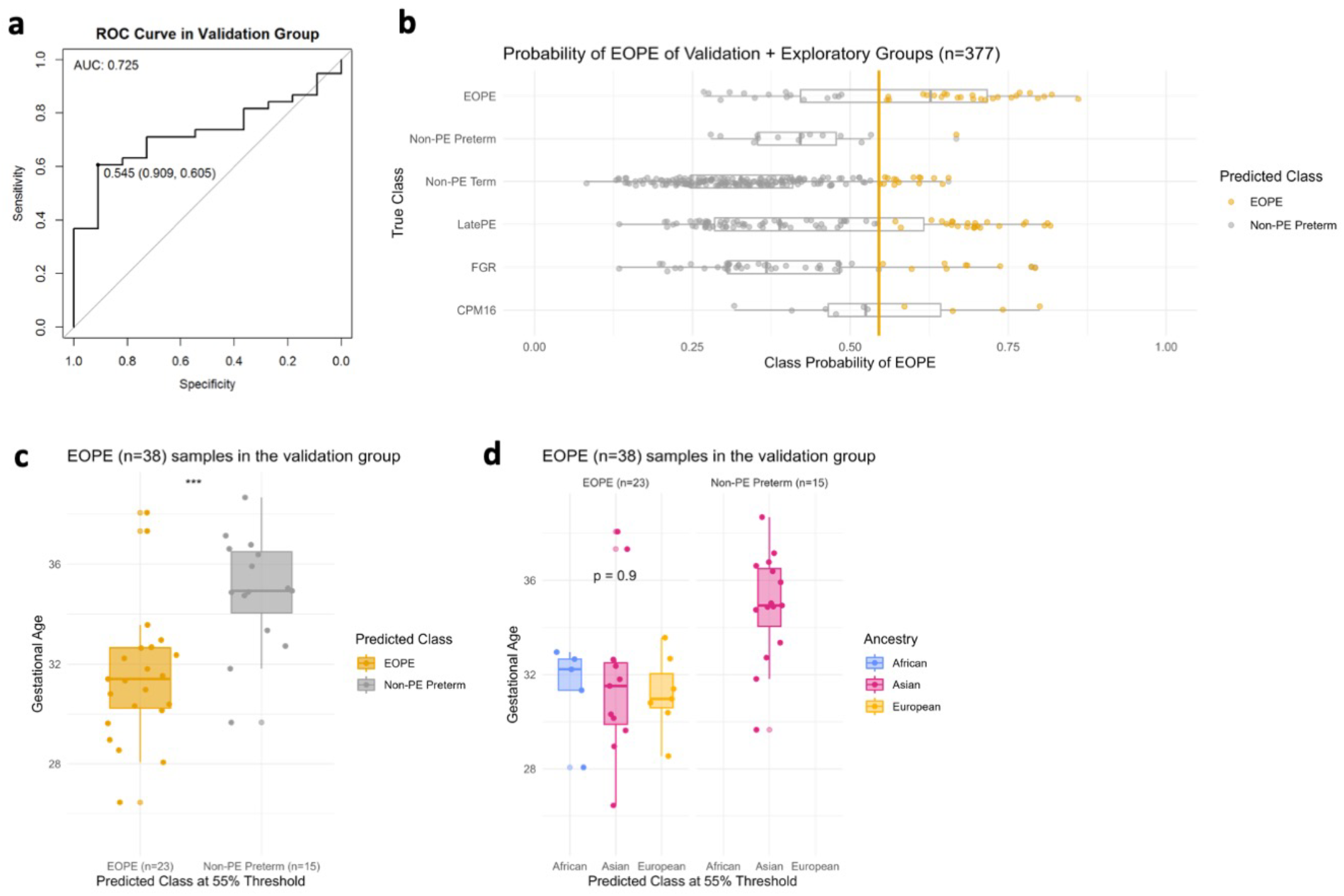
Probability of EOPE in the validation group (n=48) and in the exploratory group (n=328). eoPred was used to predict the outcome of BMIQ normalized samples in the validation and exploratory groups (the two groups were normalized separately). There are 11 nPTB samples, and 38 EOPE samples in the validation group. **a.** Receiver operating characteristic (ROC) curve in the validation group. A classification threshold of 55% was selected to maximize Youden’s J statistic (sensitivity + specificity – 1). The diagonal line indicates the curve for a classifier that predicts the majority class in all cases. **b.** Probability of EOPE of samples in the validation and exploratory groups. EOPE and nPTB samples in the plot belong to the validation group. **c**. Gestational age of EOPE samples in the validation group. eoPred predicted 23 samples as EOPE and 15 as nPTB, using a 55% probability of EOPE as the classification threshold. The difference in gestational age between EOPE samples classified as EOPE and those classified as nPTB is significant. **d.** Gestational age of EOPE samples in the validation group, faceted by ancestry groups. Ancestry was assigned to samples using planet probabilities. The probability of EOPE is not significantly different across ancestries in those samples correctly classified as EOPE.

Of the 38 EOPE samples in the validation group, 23 were predicted as EOPE (60% sensitivity) with a mean EOPE probability of 70% (Figure 2). the EOPE samples that were correctly classified as EOPE (31.85 weeks) had significantly lower average gestational age (p=0.00037) between and those that were misclassified as nPTB (35.46 weeks) (Figure 2c). In addition, 15 EOPE samples were misclassified as nPTB, these samples were all from the same dataset (GSE125605), which consisted of samples of Han Chinese ancestry as reported by the original authors (Figure 2d). All samples from GSE98224 (16 EOPE, 5 nPTB) were correctly classified. There was a wide range of EOPE probability values in the EOPE group, ranging from 56% to 82% (Figure 2b).

In the small sample size (n=11) of nPTB cases in the validation group, eoPred had a high specificity of 91%. The probability of EOPE assigned to nPTB cases, 10 of which were correctly classified as nPTB using a 55% probability threshold, ranged from 24% to 53% (Figure 2b), with an average of 41%. The misclassified nPTB sample had a EOPE probability of 67%. Applying eoPred to the validation cohort produced an area under the curve (AUC) value of 0.725, meaning that 73% of the time, for a randomly-selected pair of nPTB and EOPE samples, the model will correctly assign a higher absolute risk to the sample with EOPE than to the sample without EOPE. Generally, an AUC of 0.7-0.8 is considered good discrimination (Applied Logistic Regression, 3rd Edition, n.d.).

Lastly, we also tested the 5 CpG model in the validation group. The predictions made by eoPred (45 CpGs) and the 5 CpG model were very similar (MAE in EOPE probability between the two models=0.4). The 5 CpG model predicted EOPE with 61% sensitivity, and correctly classified 25 EOPE samples. The 5 CpG model demonstrated 91% specificity, only misclassifying 2 more nPTB samples than eoPred (Figure S2, Supplementary). The threshold chosen as optimal using Youden’s J statistic was also remarkably similar (56%).

### FGR and late-PE cases with earlier gestational ages tend to be classified as EOPE

We then predicted the disease status of 328 samples in the exploratory group, which included 2 EOPE, 86 late-PE, 3 nPTB, 181 nTB, 46 FGR, and 10 CPM16 cases. We applied eoPred to these samples to further explore its ability to detect true negatives (normotensive samples) and to investigate the EOPE probability of cases from pathologies that may overlap with the placental phenotype of EOPE, such as FGR and late-PE.

Of cases known with high confidence to be normotensive, 91% were correctly classified as nPTB (n=184, 181 nTB and 3 nPTB), with a mean EOPE probability of 31%. The 2 EOPE cases in the exploratory group were correctly assigned a high EOPE of 73% and 86%, respectively (Figure 2b). In addition, 28% of 86 late-PE cases and 20% of 46 FGR cases were predicted as EOPE, which could reflect the subset of cases in these groups with similar molecular pathology to EOPE. Both late-PE and FGR cases classified as EOPE had significantly lower gestational ages than their counterparts predicted to be normotensive (Table 4), with late-PE cases classified as EOPE having an average GA of 35.65 weeks and nPTB classified as EOPE having an average GA of 33.37 weeks. Moreover, of the 6 late-PE cases that presented with co-occurring FGR (which is commonly associated with EOPE), 4 were classified as EOPE with a mean EOPE probability of 69%, whereas 2 were predicted to be normotensive with 54% and 28% EOPE probability, respectively. Late-PE+FGR cases were more likely to be predicted EOPE (mean p(EOPE) = 69%); but not significantly more than late-PE without FGR (mean P(EOPE)=41%, p-value=0.15).

**Table 4.**
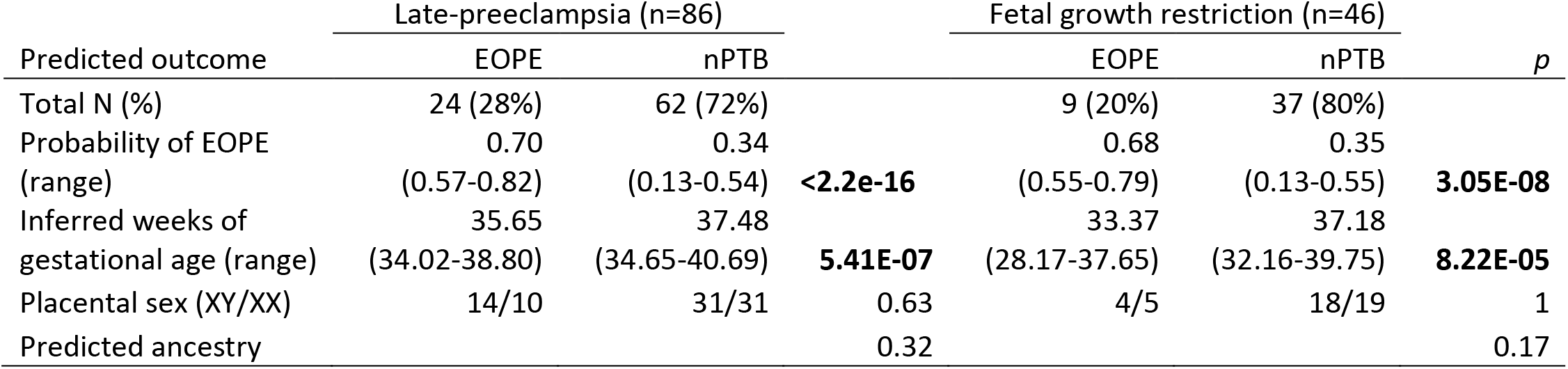
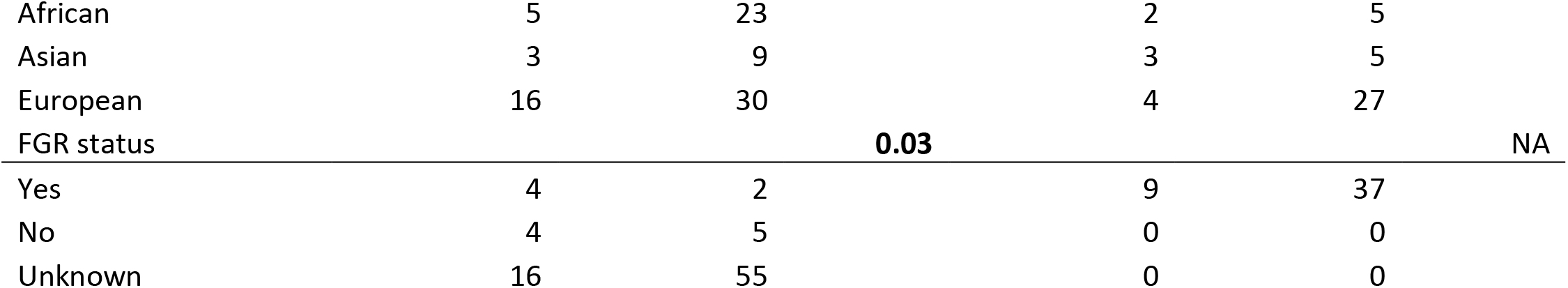
Characterization of late-PE and FGR cases in the exploratory cohort, by their predicted outcome. P-values for categorical values were calculated with Fisher’s exact test. Welch’s t-test was used to compute the p-values of the continuous variables.

Confined placental mosaicism for chromosome 16 (CPM16) is highly associated with EOPE and almost always results in FGR. DNAme patterns of CPM16 placentae were previously reported to overlap with those observed in EOPE (Yong et al., 2003). Using the same data as in (Yong et al., 2003), we evaluated how eoPred would classify CPM16 samples (n=10). Five of the 10 CPM16 samples were also diagnosed as EOPE; eoPred correctly predicted four of these samples as EOPE, while the fifth fell just below our 55% probability cut-off. All 5 CPM16 samples in which PE had not been diagnosed were correctly classified as nPTB.

### Relationship of eoPred with placental cell composition

DNAme patterns associated with PE have been reported both in cord blood and placenta, but with little overlap between the two, likely due to tissue-specificity of DNAme. We applied eoPred to 110 cord blood samples (including EOPE, LOPE, nPTB and nTB) to test whether the 45 CpG sites in our placental model could predict disease status in cord blood. However, all 110 samples were predicted as nPTB (average probability of 93%), demonstrating that the DNAme signature of EOPE developed in this study is specific to placental tissue.

Many variables have been reported to influence the placental DNA methylome throughout gestation, including cell composition, GA, and genetic ancestry. There were significant differences (Bonferroni adjusted p-value < 0.05) in the PlaNET-predicted proportion of stromal and Hofbauer cells between EOPE and nPTB (original author labels) in the training data, and in the proportion of stromal but not Hofbauer cells in the validation data (Figure S3, Supplementary Materials). However, none of the 45 predictive CpGs overlapped with any of the first or third trimester cell-type specific CpGs used by PlaNET to estimate cell composition, suggesting that these CpGs are not strongly driven by cell composition differences and should be robust to minor cell composition variation arising from sampling differences. The probability of EOPE was not strongly and significantly associated with any cell type proportion (R>0.3 and p-value<0.05) regardless of the predicted class of the samples, with the exception of cytotrophoblast, which was moderately (R=0.31, p=0.0053) associated with the probability of EOPE in samples in the validation and exploratory groups predicted as EOPE (Figure 3).

**Figure 3.**
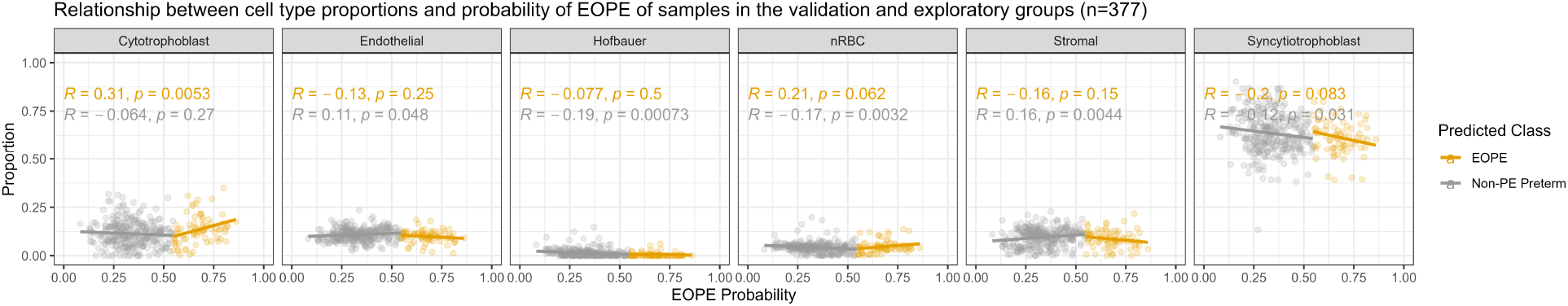
Relationship of eoPred’s EOPE probability and predictive CpG signature with placental cell types. Relationship between cell type proportions and probability of EOPE of 38 EOPE samples in the validation group. Syncytiotrophoblast and trophoblast are moderately associated with the probability of EOPE of samples misclassified as nPTB.

We also used the placental methylome browser (Yuan, 2021) to assess the cell-specific DNAme signature of the top five most predictive CpGs (cg10994126, cg26787199, cg14605117, cg10246581) in the model (i.e., those that were selected as most stable in the CV process). While DNAme varied by cell type at these CpGs, there were no consistent changes: some CpGs had higher DNAme in trophoblast cells than other cell types, whereas Hofbauer cells had the highest DNAme at others eoPred CpGs (Figure S4, Supplementary).

### Influence of genetic ancestry on the DNAme signature of EOPE

Ancestry probabilities computed with PlaNET indicated that 65% of samples in the training group and 61% in the validation and exploratory groups were primarily of European ancestry (>75% probability) (Figure S5, Supplementary). We measured the strength of association between the probability of EOPE and each of the three ancestry probabilities using Spearman’s rank order correlation for all samples in the validation and exploratory cohorts. None of the three PlaNET ancestry probabilities (African/Asian/European) were strongly associated with the probability of EOPE (ρ < 0.2).

We then compared how the β values at the 45 predictive CpGs varied by ancestry within the EOPE and nPTB groups. DNAme at 96% of the 45 eoPred CpGs was not significantly different by ancestry within 52 samples originally labelled as nPTB in the training and validation groups, suggesting that these CpGs are not systematically affected by genetic ancestry (Figure S6, Supplementary). Interestingly, the differences at these 45 CpGs across ancestries within the eoPred-predicted groups were smaller than within the groups created based on clinical labels; for instance, only 11 CpGs showed significant differences by ancestry within samples with >55% probability of EOPE in the validation and exploratory groups (Figure S7, Supplementary), as opposed to 35 within samples defined as EOPE by dataset authors in the training and validation groups (Figure S6, Supplementary).

Lastly, we explored how eoPred’s predictions may be affected by ancestry by assessing the average DNAme β values of EOPE versus nPTB samples in the training and validation cohorts at the 45 predictive CpGs (Table 5). DNAme was, as expected, significantly lower in EOPE compared to nPTB samples (p-value < 0.05) within each ancestry group, and within each of the datasets included in the training and validation groups, with the exception of GSE125605 (Table 5). The mean β of EOPE samples in this dataset, which is comprised exclusively of samples of Asian ancestry, was also higher than all the other datasets in both the training and validation groups (Table 4). In the training group, the mean β of EOPE samples of Asian ancestry was very similar to those of European ancestry (Table 4), and the mean β value of the 4 EOPE samples of Asian ancestry from GSE98224 was also similar (0.40). Therefore, the misclassification of a greater proportion of EOPE samples in GSE125605 compared to GSE98224 may be due to a dataset effect rather than to genetic ancestry. We confirmed that there were no major cell composition differences due to sampling between GSE125605 and GSE98224, which could have been a potential source for this dataset effect (Figure S8, Supplementary).

**Table 5.** Average β values at 45 predictive CpGs, by dataset and by ancestry. The mean β at the 45 predictive CpGs was computed for EOPE and nPTB samples separately, using author-assigned labels. Δβ is defined as the absolute difference between EOPE mean β and nPTB mean β. p-values were computed with an unpaired t-test.

| | EOPE mean $\beta$ (N) | nPTB mean $\beta$ (N) | $\Delta\beta$ | p-value |
| --- | --- | --- | --- | --- |
| **Training group (N=83)** |  |  |  |  |
| All samples | 0.47 (42) | 0.61 (41) | 0.14 | < 2.2e-16 |
| **By inferred ancestry** |  |  |  |  |
| Asian ancestry | 0.49 (7) | 0.62 (6) | 0.13 | < 2.2e-16 |
| African ancestry | 0.48 (7) | 0.62 (5) | 0.14 | < 2.2e-16 |
| European ancestry | 0.48 (28) | 0.62 (30) | 0.14 | < 2.2e-16 |
| **By dataset** |  |  |  |  |
| GSE100197 | 0.48 (22) | 0.62 (28) | 0.14 | < 2.2e-16 |
| GSE103253 | 0.47 (11) | 0.61 (8) | 0.15 | < 2.2e-16 |
| GSE57767 | 0.52 (5) | 0.59 (3) | 0.07 | 0.000408 |
| GSE73375 | 0.38 (4) | 0.48 (2) | 0.1 | 3.57E-06 |
| **Validation (N=49)** |  |  |  |  |
| All samples | 0.49 (38) | 0.55 (11) | 0.06 | 5.73E-10 |
| **By inferred ancestry** |  |  |  |  |
| Asian ancestry | 0.51 (26) | 0.55 (2) | 0.04 | 0.06439 |
| African ancestry | 0.43 (5) | 0 | NA | NA |
| European ancestry | 0.43 (7) | 0.55 (9) | 0.12 | < 2.2e-16 |
| **By dataset** |  |  |  |  |
| GSE98224 | 0.42 (16) | 0.56 (5) | 0.14 | < 2.2e-16 |
| GSE125605 | 0.53 (22) | 0.54 (1) | 0.006 | 0.8235 |
| GSE203396 | 0 | 0.53 (5) | NA | NA |

### Model performance varies with different DNAme normalization methods

As the performance of machine learning models can be affected by data transformations, we evaluated the performance of eoPred with different normalization methods using differently normalized iterations of a subset of the validation group data with IDATs available (GSE98224 and GSE125605, n=44). We assessed eoPred’s predictions on data normalized using 7 common methods: beta-mixture quantile normalization (BMIQ) (Teschendorff et al., 2013), quantile normalization (QN) (Aryee et al., 2014), subset-quantile within array normalization (SWAN) (Maksimovic et al., 2012), Dasen (Pidsley et al., 2013), noob (Triche et al., 2013), QN + BMIQ, and noob + BMIQ.

While our model was trained on BMIQ normalized data, it performed better on other iterations with different normalization methods (Table 5). Applying eoPred to quantile-normalized data demonstrated the best performance across all metrics evaluated (Table 5). This evaluation is strictly based on machine learning evaluation metrics and does not consider model interpretability. Our sample size is small, and we cannot conclude a given normalization method will consistently outperform others: testing on more data is required to systematically assess the effect of normalization on eoPred’s performance.

**Table 5.**
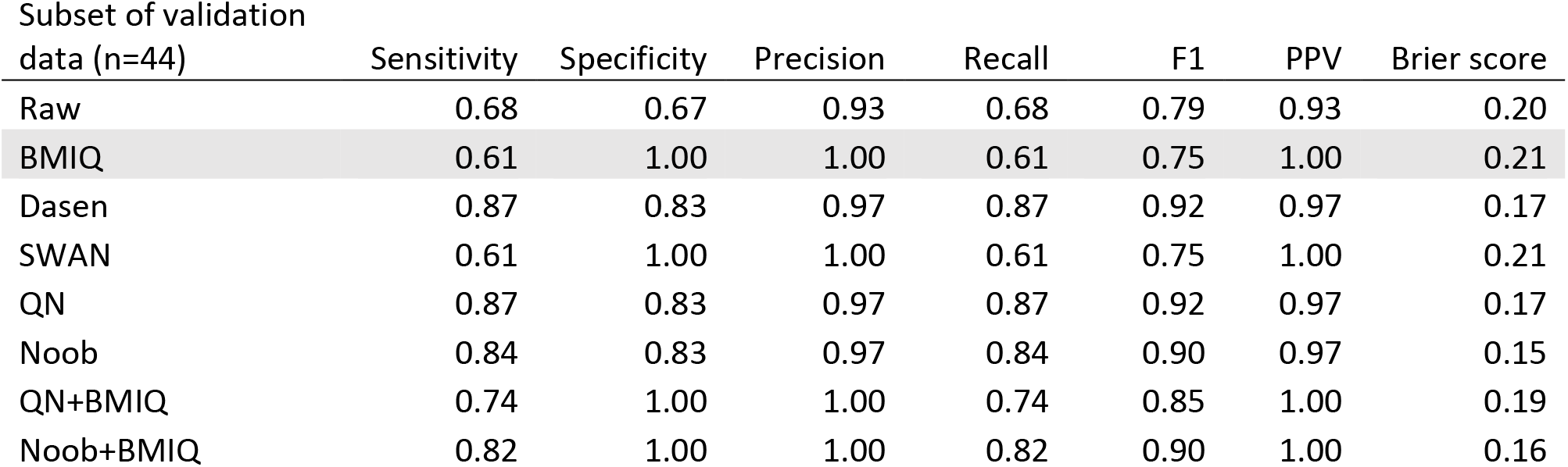
Performance metrics of eoPred’s predictions on the validation cohort, with different data transformations. Metrics of performance that resulted from prediction on filtered, BMIQ and batch corrected data are highlighted since eoPred performed best (albeit not significantly) using this processing. BMIQ (beta-mixture quantile normalization) is a within-array normalization method that corrects the methylation distribution of type II features to make them comparable to the distribution of type I features. SWAN (subset-quantile within-array normalization) is similar to BMIQ in that it adjusts the intensity distributions between the two probe types, but it does so with raw intensities. Dasen performs background adjustment followed by between-array normalization performed separately by probe design. Noob also performs background correction and dye-bias equalization. QN (quantile normalization) also performs between-array normalization.

### DNAme signature of EOPE is not strongly associated with gene expression, but may be involved in pathways relevant to placentation and preeclampsia

We next sought to investigate the biological underpinnings of the eoPred CpGs. DNAme is an important epigenetic regulator of gene expression that can affect the interactions of chromatin-binding proteins and transcription factors with DNA at the specific locus where the methyl group is present, and over moderate genomic distances (Jones, 2012). Based on the knowledge that proximal CpGs have correlated DNAme states, we grouped eoPred CpGs with proximal correlated CpG sites not included in eoPred, across samples in the training group. This was achieved by discovering 57,666 co-methylated regions (CMRs) in the training group and identifying the CMRs that comprised one or more eoPred CpGs. Only 19 of the 45 predictive CpGs occurred in CMRs; the 19 CpGs mapped to 16 distinct CMRs, which varied in size from 2 (smallest CMR) to 11 (largest CMR) CpGs. For the next step, we considered all CpGs that mapped to CMRs overlapping the 45 eoPred CpGs (n=79).

We then analyzed the EOPE versus nPTB gene expression of 14 genes that mapped to the 79 CpGs, using a linear model. Of these 14 genes, 7 were differentially expressed by EOPE status in the discovery cohort (absolute fold-change (FC)>1.4 and p-value<0.05) ; only 2 were validated with the same thresholds in an independent cohort: synapse defective rho GTPase homolog 1 (*SYDE1*) on chromosome 19, which was higher expressed in EOPE samples (FC = 1.5) and filamin B (*FLNB*) on chromosome 3, which was also more highly expressed in EOPE (FC = 1.7).

We also tested whether gene expression was significantly different between EOPE and nPTB samples for the 25 genes that the full set of 45 predictive CpGs mapped to. These 45 CpGs are distributed throughout the autosomes; 7 of them (15%) are located on chromosome 1, followed by 5 (11%) on chromosome 16 and 5 (11%) on chromosome 3. Ten genes were significantly differentially expressed in the discovery cohort by EOPE status (absolute FC>1.4 and p-value<0.05), and three of these ten genes (*SYDE1*, *FLNB*, and *SCARB1*) were differentially expressed using the same thresholds in the validation cohort. All three were higher expressed in EOPE compared to nPTB samples. Notably, pregnancy-associated plasma protein 2 (*PAPPA2*), which has been extensively reported to be associated with PE and overlaps with 3 of the 45 predictive CpGs, was identified as a DEG in the validation but not the discovery cohort.

The 45 predictive CpGs were not significantly enriched for any GO terms (FDR<0.05). The top GO term (p-value=0.0002), despite not reaching FDR significance, was “adhesion of symbiont to host”. When GO enrichment was conducted only on the 10 CpGs that map to promoter regions, “regulation of vascular associated smooth muscle cell proliferation” is the top term, though not significant after FDR multiple test correction. Lastly, we assessed whether the 45 predictive CpG probes were enriched in placentally-relevant gene sets reported in a recent study (Naismith and Cox, 2021). Once again, no gene sets were significant (FDR<0.05), but the top gene set by nominal p-value was “genes upregulated in invasive cytotrophoblasts versus human chorionic trophoblast progenitor cells”, and the genes associated with that set were *PPARG*, *PAPPA2*, and *RRBP1*, all of which were identified as differentially expressed in the discovery cohort, though none were validated.

Lastly, we evaluated whether the 45 CpGs overlapped lists of CpGs with PE-associated placental DNAme found in eleven previous studies (Martin et al., 2015; Yeung et al., 2016; Berg et al., 2017; Kim et al., 2017; Zhao et al., 2017; Wilson et al., 2018; Wang et al., 2019a; Lim et al., 2020; van den Berg et al., 2020; Workalemahu Tsegaselassie et al., 2020). Of the 45 eoPred CpGs, 36 overlapped hits found to be EOPE-associated by Wilson et al., 2018, which is to be expected given that the discovery dataset (GSE100197) was included in our training group. Of the eoPred CpGs reported in Wilson et al., cg10246581, one of the top 5 most predictive eoPred CpGs, was also reported in Wang et al., 2019 and Kim et al., 2017. The data used by Wang et al., 2019 was in our validation group (GSE125605). Another CpG among the 5 most predictive (cg26625897) was also reported to be associated with EOPE in placental tissue by 3 studies (Yeung et al., 2016; Kim et al., 2017; Lim et al., 2020).

## Discussion

Previous studies of placental DNAme have shown that placentae from pregnancies affected by preeclampsia exhibit DNA methylome differences as compared to placentae from uncomplicated pregnancies (Blair et al., 2013; Yeung et al., 2016; Leavey et al., 2018; Wilson et al., 2018). We used placental DNAme to develop a supervised model, “eoPred”, to classify placentae according to their DNAme patterns into those with and without EOPE. This tool can be applied by users to define homogeneous subgroups of placentae with an EOPE-associated DNAme phenotypes, rather than relying on clinical measures for group definition.

We anticipate that one of the major values of eoPred will be that it can be used to identify placentae from various pregnancy complications that share a similar placental phenotype with EOPE, even if the pregnant parent was not diagnosed with PE. Pregnancy complications such as FGR or preterm birth, as well as environmental conditions during gestation (i.e., smoking, stress) are associated with an increased risk of developing PE (Lisonkova and Joseph, 2013; Bartsch et al., 2016); we hypothesize that eoPred will be helpful in investigating the extent to which such exposures affect the placental DNA methylome at different degrees and timepoints in gestation. Additionally, eoPred will enable the deeper investigation of DNAme datasets with limited clinical information on EOPE status, but which may have other valuable information recorded, such as drug or environmental exposure data. This will broaden the scope of available studies beyond current analysis limitations of clinically-confirmed and reported cases of PE and hopefully provide more insight into the pathogenesis of PE. Lastly, eoPred has the potential to generate translational impact in the development of early-diagnostic tools based on cell-free placental DNA methylation, in that if placental DNAme signatures can be measured in maternal blood, PE status may be identifiable from such marks (Kwak et al., 2020; Palei, 2021; Moufarrej et al., 2022).

Our model, relying on 45 predictive CpGs, was able to successfully label 60% of 38 EOPE cases and 91% of 11 nPTB cases in an independent validation group (n=49). The relatively low sensitivity of 60% primarily arose from one of the two datasets in the validation group. Without further independent clinical data, we cannot be sure that diagnostic criteria were identical in each dataset. Moreover, the significant association of the probability of EOPE with disease biology (e.g., co-occurring FGR) and gestational age suggests that the 45 CpG signature is important for the placental phenotype of EOPE. The gestational age (GA) of correctly classified EOPE samples (n=23) is significantly lower (p-value<0.05) than that of the misclassified samples (n=15). This highlights the heterogeneity that exists within preeclampsia subtypes that are defined based on GA thresholds, and suggests that, at least in terms of placental pathology and corresponding molecular changes, preeclampsia exists on a continuum of severity. In addition, this observation confirms the findings from Wilson et al. (Wilson et al., 2018) who previously observed the similarity in global DNAme patterns between samples with co-occurring late-PE and FGR diagnosed between 34 and 36 weeks, and EOPE (diagnosed prior to 34 weeks). Further, while eoPred was trained on EOPE and normotensive preterm (nPTB) samples, it is expected that normotensive term (nTB) samples will be assigned a higher probability of nPTB than of EOPE.

In validating our model, all samples in one of our validation datasets (GSE98224) were correctly classified. In contrast, in the other validation dataset (GSE125605), which is comprised exclusively of samples of Asian ancestry, 15/22 EOPE samples were misclassified as nPTB. Notably, the 7 samples in this dataset which were correctly predicted as EOPE had a significantly lower GA than those predicted as nPTB, which may suggest a biological explanation for these misclassifications. The interpretation of this finding is currently limited by the fact that all 15 misclassified samples are equally affected by dataset, ancestry, and gestational age confounding effects. Our finding that 1) the 45 predictive CpGs do not vary by ancestry within normotensive preterm samples and 2) there is a significant difference in β values at the 45 predictive CpGs between EOPE and nPTB samples in samples of Asian ancestry from datasets other than GSE125605, both in the training and validation groups, suggests that the high misclassification rate in this dataset may not be a result of ancestry but rather a dataset effect (e.g., differences in how cases were ascertained and classified as EOPE or systematic differences in sample and array processing). Moreover, the minimal differences observed by ancestry at the 45 CpGs within each eoPred-defined groups (EOPE/nPTB) suggests that if subtypes are more homogeneous when defined based on placental phenotype rather than clinically, they may be more robust to genetic variation differences. European ancestry is overrepresented in a majority of placental DNAme datasets, and more genetically diverse data is needed to appropriately assess the utility of eoPred in different populations. Ideally, a new iteration of the tool can be built with a larger and more diverse sample size to further investigate this.

While more validation is needed, many of the CpGs selected as predictive by our model are relevant to preeclampsia. For example, there were 3 eoPred CpGs in the *PAPPA*-2 gene in chromosome 1, which were some of most stable features in our model. *PAPPA*-2 is highly expressed in the placenta (Wang et al., 2009; Sifakis et al., 2018) and has been reported to be upregulated in cases of severe PE (Macintire et al., 2014; Kramer et al., 2016), although it was not validated as differentially expressed between EOPE and nPTB samples in our study. In addition, two genes that two of the predictive CpGs mapped to and that were significantly differentially expressed in our independent cohort, *SYDE1* and *FLNB*, have also been reported as potentially involved in FGR and PE in some studies (Tejera et al., 2013; Lo et al., 2017; Leavey et al., 2018; Huang et al., 2022). In addition, the model demonstrated a high specificity in both validation and exploratory groups, where 171 of 195 normotensive samples were predicted correctly, adding confidence to the utility of eoPred.

The performance of machine learning models on new data may be affected by data transformations (e.g., normalization), particularly if these differ from the transformations that were applied to the data that the model was trained on. Interestingly, our results using our validation group suggest that the prediction ability of eoPred is improved with quantile normalization, while our training data was normalized using BMIQ. There are two possible interpretations for this result. Since gestational age was lower in EOPE samples classified as EOPE than in those classified as nPTB, it is possible that EOPE samples with a low EOPE probability have a milder placental phenotype that is more similar to that of late-onset preeclampsia. However, given the overrepresentation of samples from GSE125605 among the misclassifications, this deserves further study. The better performance of eoPred on quantile normalized as opposed to BMIQ normalized data could also be explained by the fact that quantile normalization is a between-array method, which means it considers all samples in the data, and transforms the data to minimize global DNAme differences between samples (Hicks et al., 2018). As such, quantile normalization could be correcting for dataset-associated differences in a similar way to batch correction, and as such reducing noise arising from technical variation, which could improve the performance of eoPred. We encourage users to make an informed choice on what the best normalization method is in the context of their data, and if choosing a method other than BMIQ, we recommend comparing eoPred’s predictions on BMIQ-normalized data to data normalized using the method of choice, if different.

Placental DNAme from whole tissue is a composite of cell-specific DNAme patterns, and while placental cell composition often varies between cohorts due to sampling differences, it has also been proposed to vary in preeclamptic placentae for biological reasons (Yuan et al., 2021; Campbell et al., 2023). Furthermore, preeclampsia-associated differentially expressed genes in cell-free RNA were reported to be tissue- or cell-type specific (Moufarrej et al., 2022). Although EOPE samples in our training and validation groups had less predicted stromal and Hofbauer cells compared to nPTB, the magnitude of change was small, and no difference was observed for the two trophoblast subtypes, which make up the bulk of the placental (chorionic villus) sample. Furthermore, DNAme variation at the 45 EOPE predictive CpGs did not appear to be driven by cell composition. We observed that the probability of EOPE was moderately associated with increased cytotrophoblast estimates particularly within samples predicted as EOPE in the validation and exploratory groups, but not at any of the other cell types. Overall, the EOPE-associated DNAme signature used by eoPred does not seem to be significantly influenced by cell composition.

There are several strengths and limitations inherent to how we have constructed eoPred given the available data and how it was clinically characterized. The Illumina HM450K array is the most common platforms used for placental studies of PE, and eight Illumina HM450K publicly available on GEO were used to train, validate, and explore the utility of our model. Our use of the HM450K platform in this study will increase eoPred’s relevance to future EWAS studies, as will our choice to train the model on CpGs that are also present on the EPIC array, which will enable the use of eoPred with new datasets as they are generated. While the HM450K array only measures DNA methylation at ∼1% of all CpGs in the genome, there are widespread changes in PE at several regions covered by this array. Secondly, re-utilization of public data is essential to maximize research time and resources, but it comes with limitations when the purpose is to ask new questions. In compliance with privacy and confidentiality agreements, public data often lacks extensive clinical and/or phenotypical characterization of samples. This limited our ability to interpret our findings; in particular, we would like to investigate whether the probability of EOPE is associated with severity of clinical measures and/or placental histopathology, and whether the placental samples from other reported etiologies with high EOPE probability exhibited characteristic pathological features also seen in EOPE. More generally, public datasets of chorionic villi samples of various etiologies are sparse, which limited the design of our predictor. As suggested by Myatt et al. (Myatt Leslie et al., 2014), comparing PE to “normal” outcomes will inevitably lead to falsely predictive capability, begging the question of whether PE will be adequately identified as distinct from other outcomes, both normal and abnormal. While this is certainly a limitation of our study, we chose a binary classification model as opposed to a multiclass model due to the size constraints of our data, and we developed eoPred as a tool with research utility in mind, with the aim that its application to future datasets may provide insight into the pathophysiology of PE in the placenta, and other associated outcomes, As our understanding of PE grows, this study may lay some groundwork for the development of future clinical models.

## Conclusion

In this study, we develop eoPred, which classifies placentae as EOPE or nPTB, and outputs a continuous measure of the placental phenotype of EOPE. This tool has been included in the PlaNET R package and is available at https://www.bioconductor.org/packages/release/bioc/html/planet.html. We anticipate that eoPred will be useful in future EWAS studies to measure the influence of EOPE on the placental DNA methylome, and to study the effect of other variables of interest such as maternal exposures, chromosomal sex, and stress on a molecularly homogeneous subtype of preeclampsia. Importantly, we confirmed that this measure of EOPE probability is robust to cell composition differences that can arise from sampling and/or pathology, and that it works adequately with a variety of DNAme normalization methods. Lastly, the 45 CpG DNAme signature of EOPE appears to be shared with other pregnancy complications that have some overlapping clinical and placental phenotypes with EOPE, illustrating the overlap between pregnancy complications mediated by the placenta.

## Declarations

### Ethics approval and consent to participate

Ethics approval for use of human research subjects in this study was obtained from both the University of British Columbia and BC Women’s and Children’s Hospital ethics committees in Vancouver, BC, Canada (H04–704488). Informed written consent was obtained from all study participants.

### Consent for publication

Not applicable.

### Availability of data and materials

eoPred is available as a function in the PlaNET R package (https://www.bioconductor.org/packages/release/bioc/html/planet.html). All datasets used in this study are publicly available via the Gene Expression Omnibus, with dataset accession numbers GSE100197, GSE98224, GSE125605, GSE110829, GSE73375, GSE75196, GSE57767, GSE103253 and GSE120981. The 45 EOPE-specific CpGs selected in our model can be found in Table S4. Source code and R Markdown scripts with the processing and analysis pipeline can be found at https://github.com/iciarfernandez/eoPred/.

## Data Availability

eoPred is available as a function in the planet R package (https://www.bioconductor.org/packages/release/bioc/html/planet.html). All datasets used in this study are publicly available via the Gene Expression Omnibus. The 45 EOPE-specific CpGs selected in our model can be found in Table S4. Source code and R Markdown scripts with the processing and analysis pipeline can be found at https://github.com/iciarfernandez/eoPred/.

## Competing interests

The authors declare that they have no competing interests.

## Funding

This work was supported by a Canadian Institutes of Health Research (CIHR) grant to WPR [F19-04091]. WPR holds a CIHR Research Chair in Sex and Gender Science [GSK-171375] and an investigatorship award from the BC Children’s Hospital Research Institute. IFB receives support from a UBC Four-Year Fellowship.

## Authors’ contributions

WPR, AMI, and VY contributed to project design. IFB performed analysis of data and wrote manuscript. AMI processed gene expression data. VY helped develop eoPred functions for the PlaNET package. All authors read, edited, and approved the final manuscript.

## Acknowledgements

We thank pregnant people who generously choose to donate their placentae for the advancement of research, and the scientific community for their commitment to making data publicly available. This work would not be possible without current and past members of the Robinson lab, we especially acknowledge Dr. Maria Peñaherrera A. for thoughtful discussion and feedback on the design and data analysis.

## Authors’ information

**BC Children’s Hospital Research Institute, Vancouver, BC, Canada**

Icíar Fernández Boyano, Amy M. Inkster, Victor Yuan & Wendy P. Robinson

**Department of Medical Genetics, University of British Columbia, Vancouver, BC, Canada**

Icíar Fernández Boyano, Amy M. Inkster, Victor Yuan & Wendy P. Robinson

## Notes

### Competing Interest Statement

The authors have declared no competing interest.

### Author Declarations

The University of British Columbia Children's Hospital and Women's Health Centre of British Columbia Research Ethics Boards (UBC C&W REB) gave ethical approval for this work (H04-704488).

