## Supplementary Materials for "eoPred: Predicting the placental phenotype of early-onset preeclampsia using DNA methylation"

### Figure S1

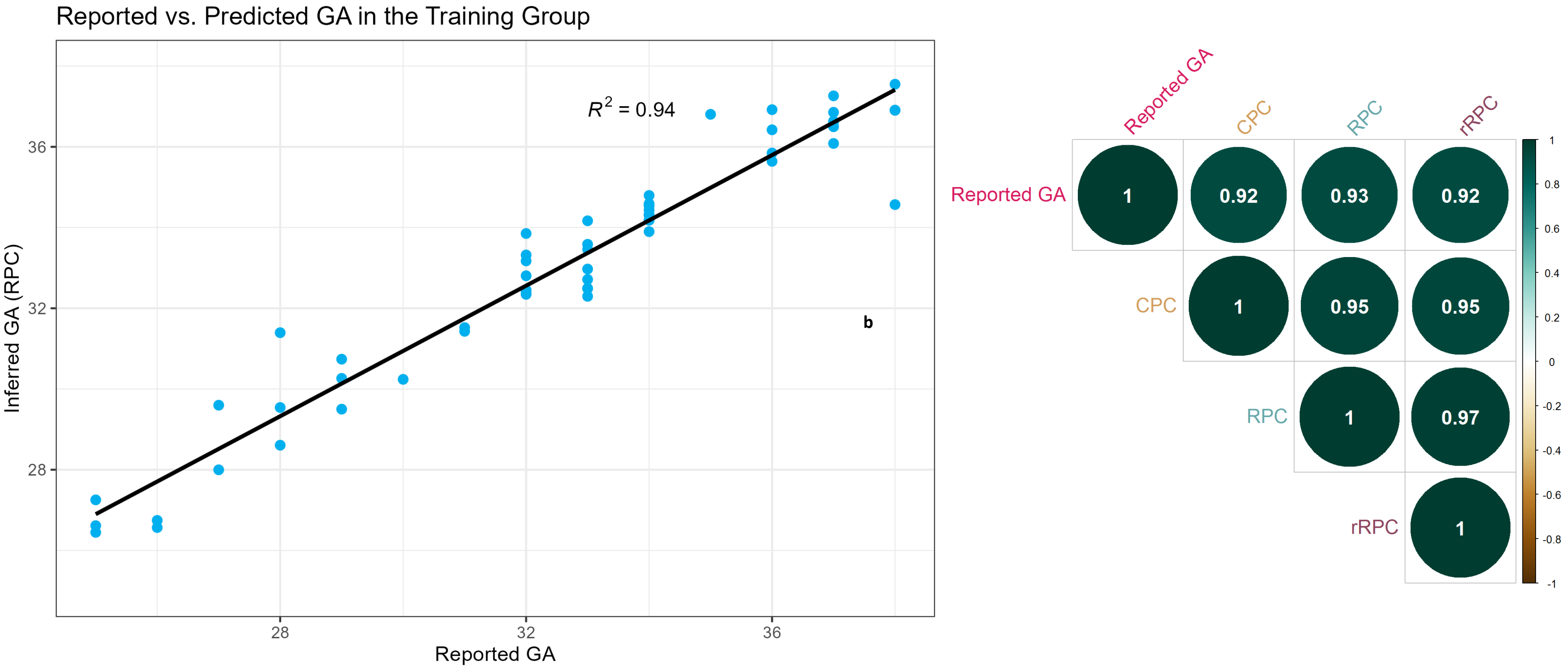


**Figure S1. Relationship between reported and inferred gestational age (GA).** Gestational age was inferred for all samples using the robust placental clock (Lee et al., 2019) as implemented in the R package planet. a. Association between reported and predicted GA in the training group, specifically in GSE100197 and GSE73375, which are the only 2 datasets out of 4 in the training group for which authors reported GA in weeks. b. Correlation between reported GA and inferred GA of all samples from GSE125605, GSE110829, GSE73375, and GSE75196, which are the only 4 datasets in this study that were not included as part of the training of the robust placental clock.

### Figure S2

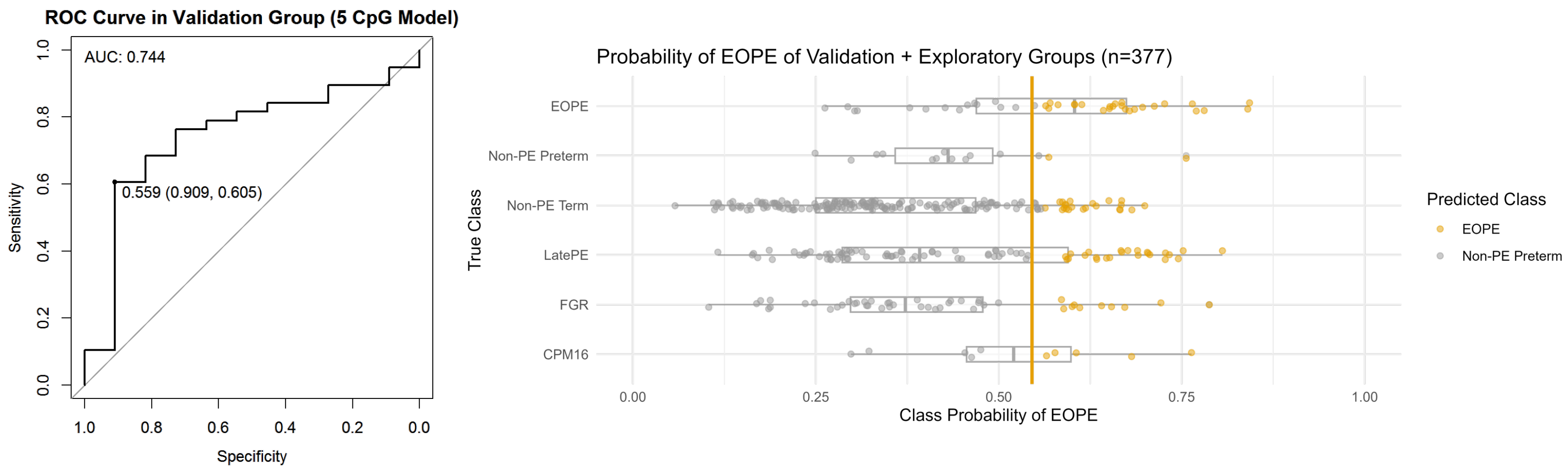


**Figure S2. Probability of EOPE using a 5 CpG model in the validation group (n=48) and in the exploratory group (n=328).** A 5 CpG model was used to predict the outcome of BMIQ normalized samples in the validation and exploratory groups (the two groups were normalized separately). There are 11 nPTB samples, and 38 EOPE samples in the validation group. **a.** Receiver operating characteristic (ROC) curve in the validation group. A classification threshold of 56% was selected to maximize Youden's J statistic (sensitivity + specificity – 1). The diagonal line indicates the curve for a classifier that predicts the majority class in all cases. **b.** Probability of EOPE of samples in the validation and exploratory groups. EOPE and nPTB samples in the plot belong to the validation group.

### Figure S3

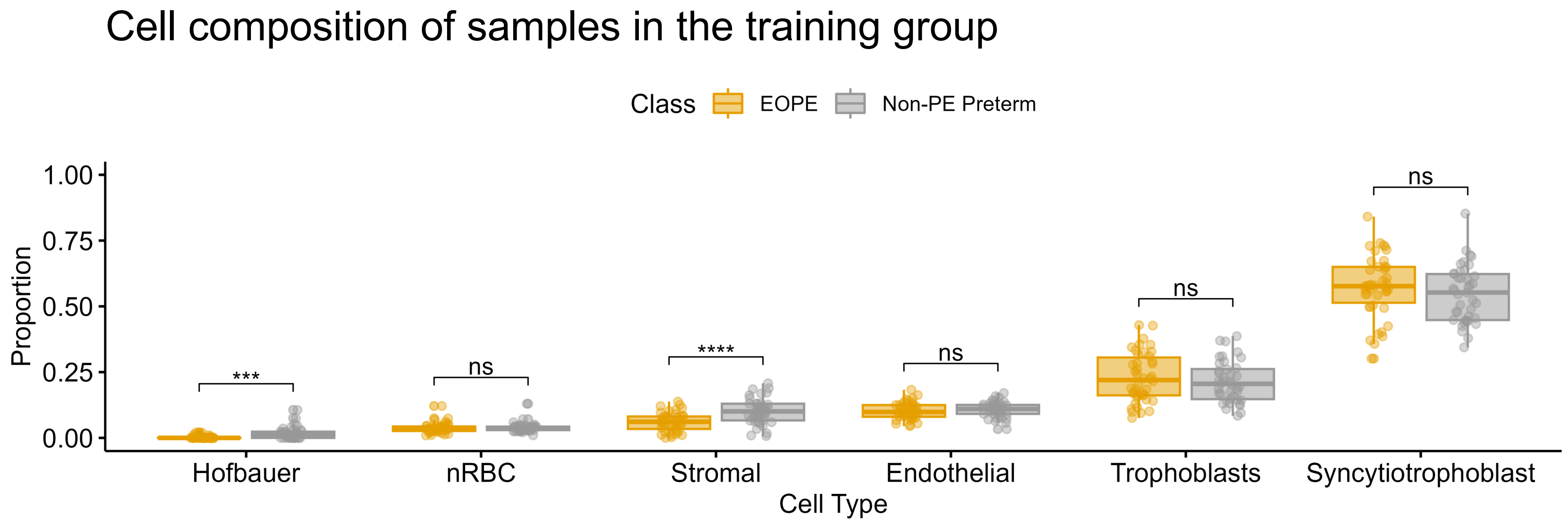


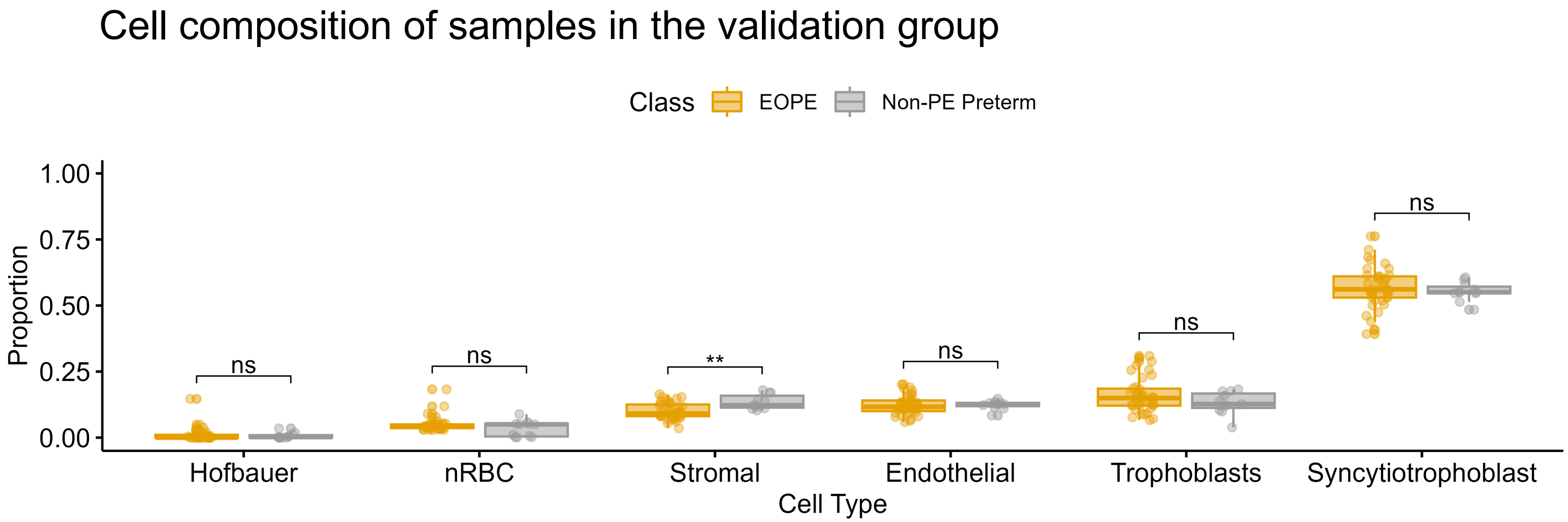


**Figure S3. Cell composition of samples in the training (n=83) and in the validation (n=48) groups.** Placental cell composition was estimated using the R package planet (Yuan et al., 2021).

### Figure S4

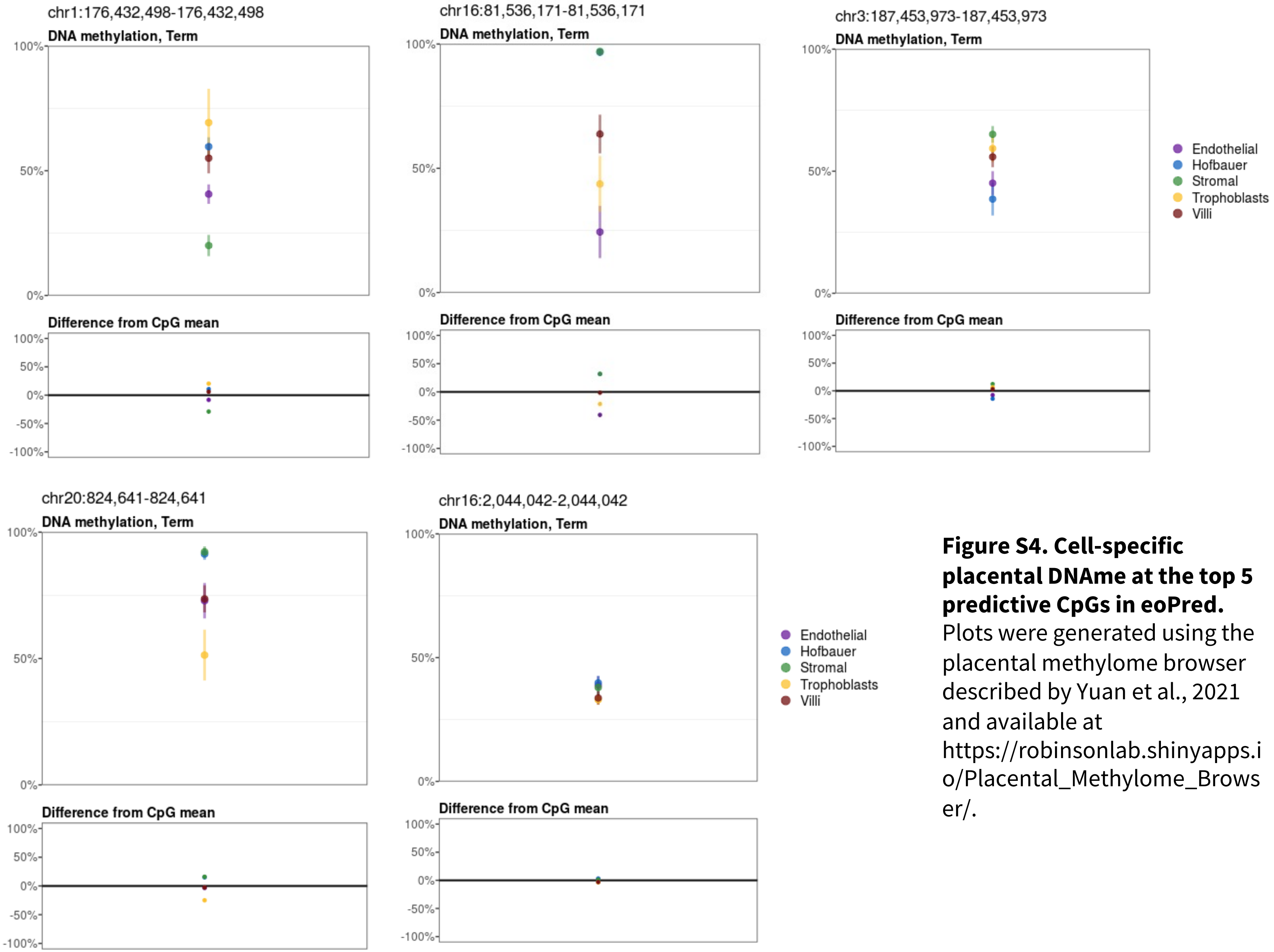


**Figure S4. Cell-specific placental DNAme at the top 5 predictive CpGs in eoPred.** Plots were generated using the placental methylome browser described by Yuan et al., 2021 and available at https://robinsonlab.shinyapps.io/Placental_Methylome_Browser/.

### Figure S5

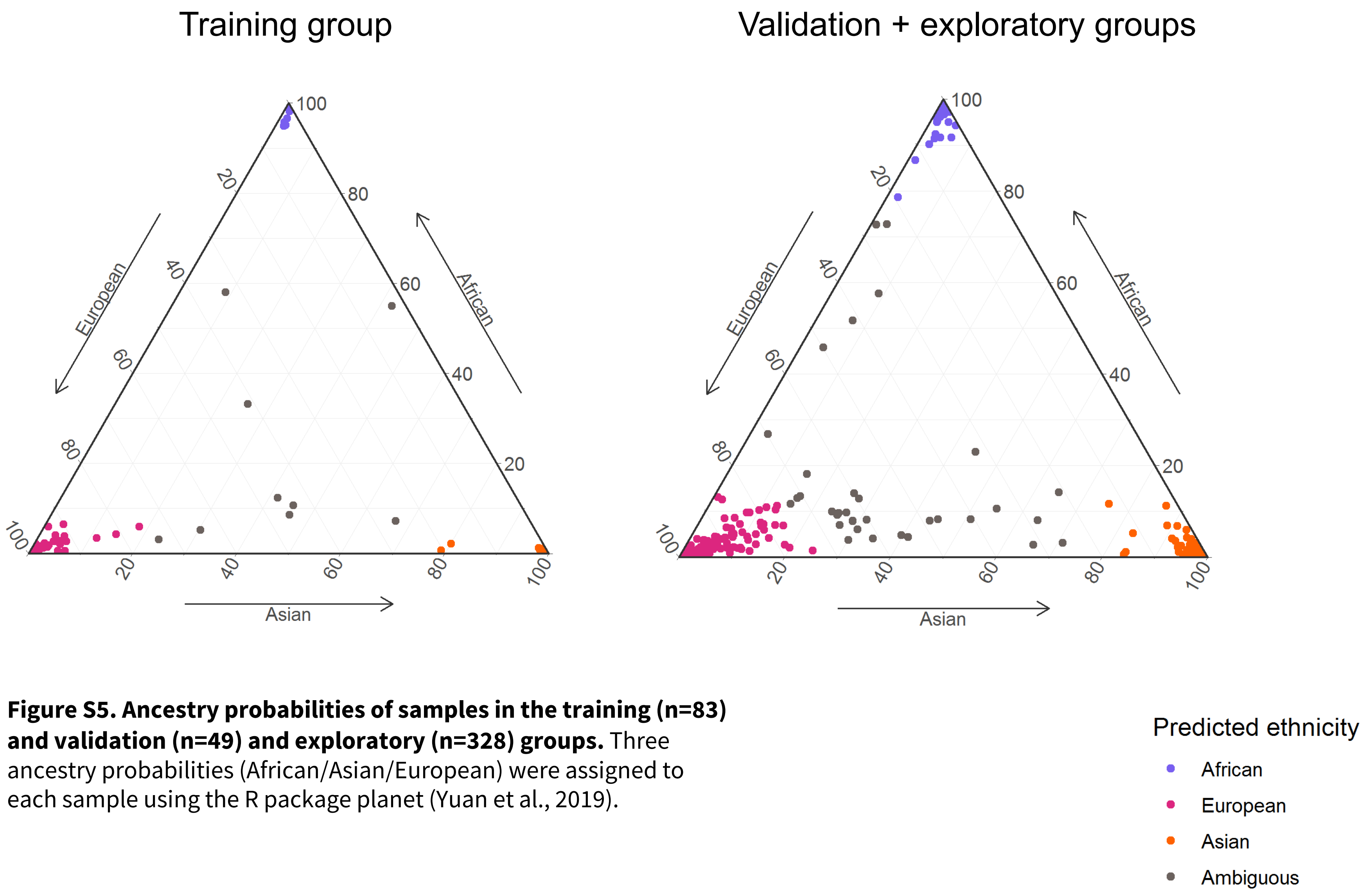


**Figure S5. Ancestry probabilities of samples in the training (n=83) and validation (n=49) and exploratory (n=328) groups.** Three ancestry probabilities (African/Asian/European) were assigned to each sample using the R package planet (Yuan et al., 2019).

### Figure S6

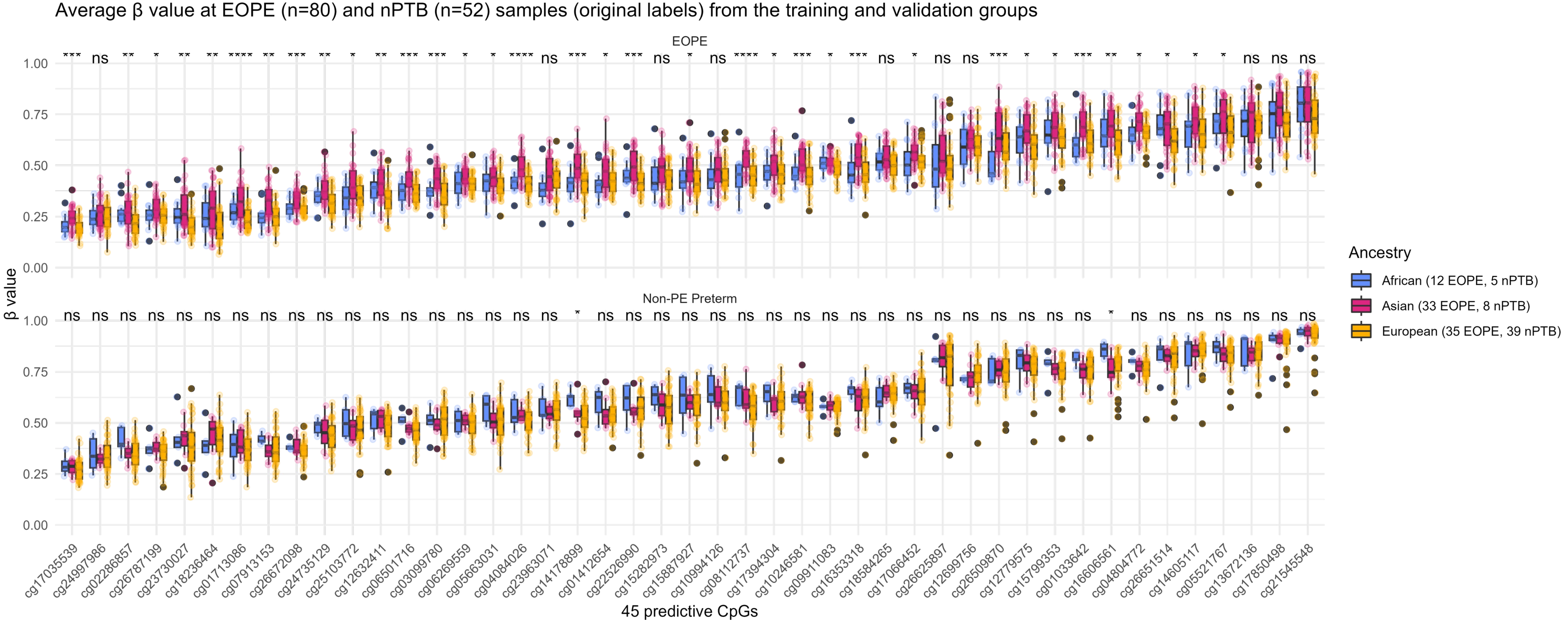


**Figure S6. Average β values at the 45 predictive CpGs used by eoPred in EOPE (n=80) and nPTB (n=52) samples in the training and validation groups.** There are 17 samples of African ancestry, 41 samples of Asian ancestry, and 74 samples of European ancestry depicted in the plot. Ancestry was inferred using three ancestry coordinates based on DNA methylation with the R package planet. EOPE and nPTB labels are based on the labelling by the original authors of each dataset and not on eoPred-assigned probabilities.

### Figure S7

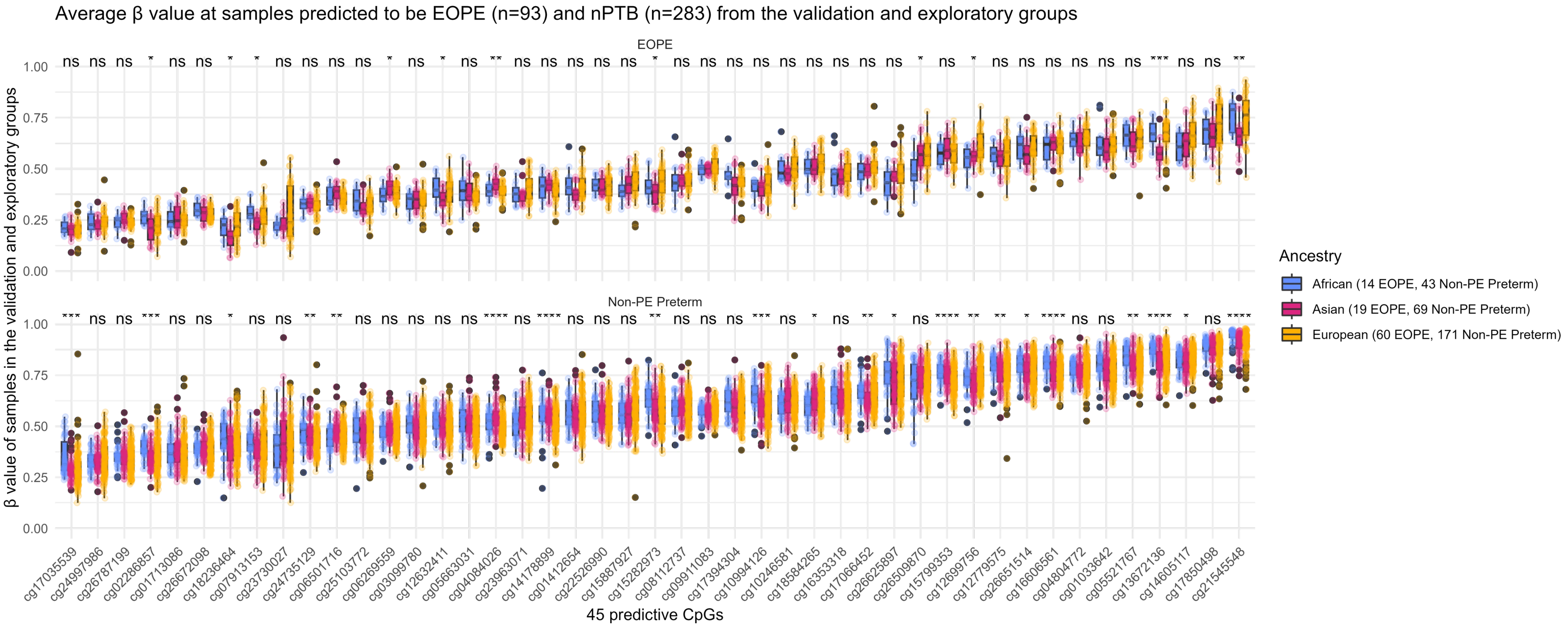


**Figure S7. Average β values at the 45 predictive CpGs used by eoPred in EOPE (n=93) and nPTB (n=283) samples in the validation and exploratory groups.** Sample groups (EOPE/nPTB) are defined based on eoPred-assigned probabilities. Samples with >55% probability of EOPE are classified as EOPE. There are 57 samples of African ancestry, 88 samples of Asian ancestry, and 231 samples of European ancestry depicted in the plot. Ancestry was inferred using three ancestry coordinates based on DNA methylation with the R package planet.

### Figure S8

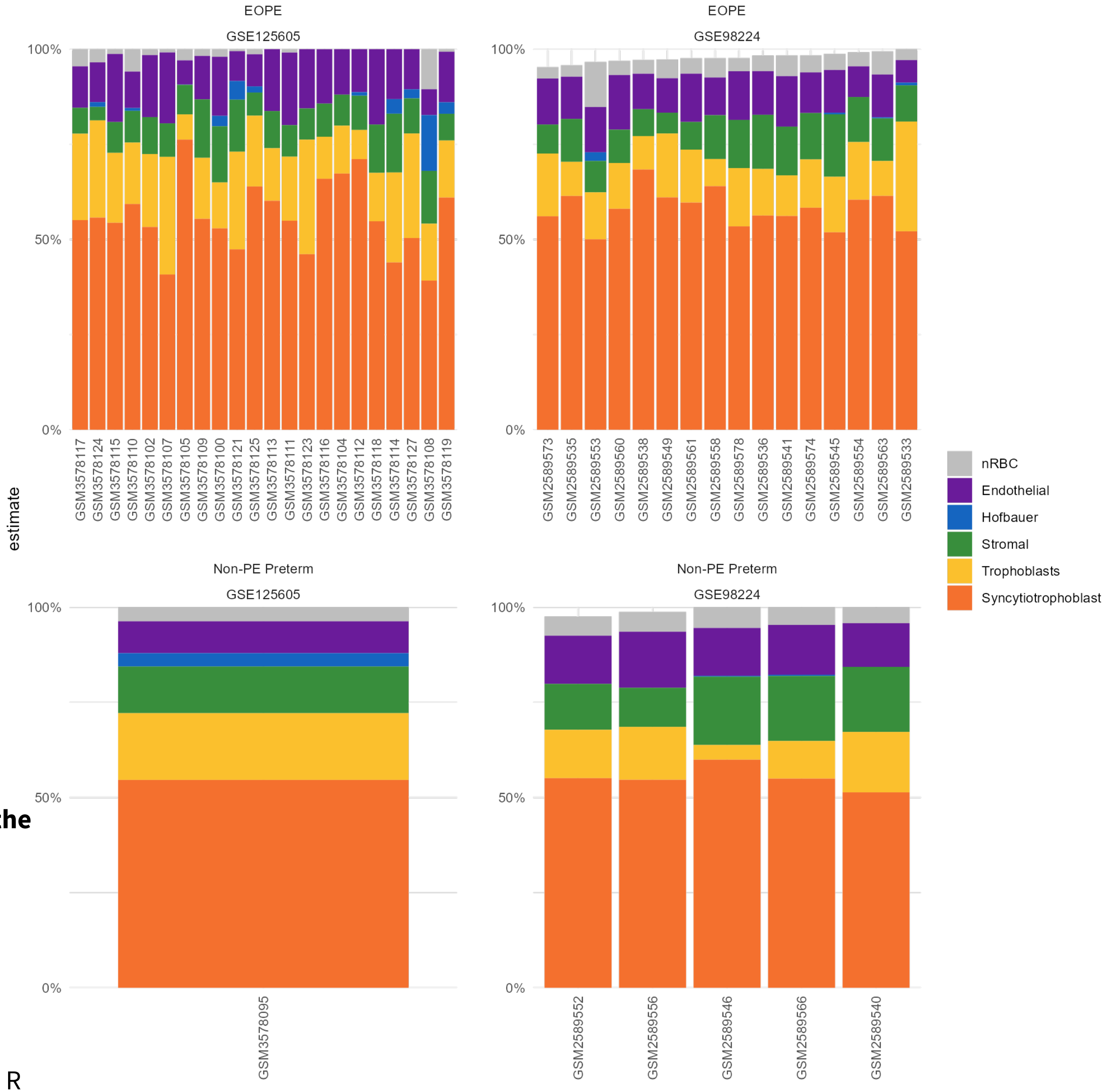


**Figure S8. Cell composition of EOPE and nPTB samples in the validation group.** Two datasets from the validation group are included in this plot (GSE125605 and GSE98224). Cell composition estimates were inferred using the R package planet.
